# The tumor multi-omic landscape of endometrial cancers developed on a germline genetic background of adiposity

**DOI:** 10.1101/2023.10.09.23296765

**Authors:** George Richenberg, Amy Francis, Carina N. Owen, Victoria Gray, Timothy Robinson, Aurélie AG Gabriel, Kate Lawrenson, Emma J. Crosbie, Joellen M. Schildkraut, James D. Mckay, Tom R. Gaunt, Caroline L. Relton, Emma E. Vincent, Siddhartha P. Kar

## Abstract

High body mass index (BMI) is a causal risk factor for endometrial cancer but the tumor molecular mechanisms affected by adiposity and their therapeutic relevance remain poorly understood. Here we characterize the tumor multi-omic landscape of endometrial cancers that have developed on a background of lifelong germline genetic exposure to elevated BMI. We built a polygenic score (PGS) for BMI in women using data on independent, genome-wide significant variants associated with adult BMI in 434,794 women. We performed germline (blood) genotype quality control and imputation on data from 354 endometrial cancer cases from The Cancer Genome Atlas (TCGA). We assigned each case in this TCGA cohort their genetically predicted life-course BMI based on the BMI PGS. Multivariable generalized linear models adjusted for age, stage, microsatellite status and genetic principal components were used to test for associations between the BMI germline PGS and endometrial cancer tumor genome-wide genomic, transcriptomic, proteomic, epigenomic and immune traits in TCGA. High BMI germline PGS was associated with (i) upregulated tumor gene expression in the *IL6*-*JAK*-*STAT3* pathway (FDR=4.2×10^−7^); (ii) increased estimated intra-tumor activated mast cell infiltration (FDR=0.008); (iii) increased single base substitution (SBS) mutational signatures 1 (FDR=0.03) and 5 (FDR=0.09) and decreased SBS13 (FDR=0.09), implicating age-related and APOBEC mutagenesis, respectively; and (iv) decreased tumor EGFR protein expression (FDR=0.07). Alterations in *IL6*-*JAK*-*STAT3* signaling gene and EGFR protein expression were, in turn, significantly associated with both overall survival and progression-free interval. Thus, we integrated germline and somatic data using a novel study design to identify associations between genetically predicted lifelong exposure to higher BMI and potentially actionable endometrial cancer tumor molecular features. These associations inform our understanding of how high BMI may influence the development and progression of this cancer, impacting endometrial tumor biology and clinical outcomes.

## INTRODUCTION

Endometrial cancer is the second most common malignancy of the female genital tract and the sixth most common cancer among women worldwide (1). In 2020, there were over 417,000 new endometrial cancer cases and 97,000 deaths from this cancer globally (1), and its incidence and mortality are rising (2). Several risk factors are associated with endometrial cancer (3-5). These include higher body mass index (BMI; the most common measure of adiposity) and obesity, long-term unopposed estrogen exposure, insulin resistance and diabetes, and inherited genetics, particularly Lynch syndrome that involves germline mutations in mismatch repair genes. Over 50% of endometrial cancer in the population is attributable to overweight and obesity (6, 7). Meta-analyses of observational studies have shown that the relative risk for endometrial cancer for a 5 kg/m^2^ increase in BMI is approximately 1.5 (8) and those in the highest BMI category have a 7-fold elevated risk of developing this cancer than those with normal BMI (9). Mendelian randomization (MR) studies (10), which leverage the random allocation at conception of germline genetic variants robustly associated with BMI to proxy BMI over the course of life have confirmed that the association between higher BMI and endometrial cancer risk is causal (11-13).

Investigating the biological basis of the adiposity-endometrial cancer link has been identified as a top research priority by patients, survivors, carers and clinicians (14). MR mediation analyses have suggested that only 7 to 19% of the effect of BMI on endometrial cancer risk may be mediated by circulating insulin, testosterone and sex hormone binding globulin levels (11), indicating that most of the effect of BMI occurs via mechanisms that are as yet unexplained. The search for endometrial tissue-specific effects of high BMI has been elusive and focused on the interface between IGF1 receptor signaling and the PI3K/AKT/mTOR pathway in the context of endometrial hyperplasia (15-17). These studies have been limited by the use of non-human model organisms, and, for human studies, small sample sizes and an emphasis on cell line experiments. Moreover, despite the established association between increased BMI and endometrial cancer, little is known about how long-term adiposity shapes endometrial tumor biology. Relationships between long-term adiposity and the endometrial tumor molecular profile may reflect key steps in tumor development and progression and thereby reveal actionable preventative and therapeutic targets. Such relationships have the potential to form the basis of precision oncologic interventions in women with a history of overweight or obesity who develop endometrial cancer.

Here we apply a unique study design that uses a polygenic score (PGS) for BMI to evaluate the association between adiposity and endometrial cancer tumor multi-omic traits. BMI PGS, which represents the aggregate effects of multiple BMI-associated germline variants, better captures lifelong trajectories of adiposity (18, 19). In contrast, BMI measured at a single timepoint such as at cancer diagnosis may be affected by the developing cancer itself (reverse causation) or confounded by other factors. We exploit The Cancer Genome Atlas (TCGA) Uterine Corpus Endometrial Carcinoma (UCEC) project data that includes tumor genomic, transcriptomic, proteomic and epigenomic profiles of over 350 endometrial cancer tumors with matched blood-based germline genotype data, assigning each participant in this cohort their PGS for BMI (20). We obtain this BMI PGS from a genome-wide association study (GWAS) of over 434,000 women (18), combining the effects of independent, genome-wide significant single nucleotide polymorphisms (SNPs) associated with BMI. Utilizing this study design integrating BMI GWAS data with the germline and somatic components of the TCGA endometrial cancer cohort enabled us to uncover key associations between genetic liability to elevated BMI and tumor genomic, transcriptomic and proteomic features, including features with an impact on cancer survival. These associations may offer potential opportunities for early intervention and targeted treatment in endometrial tumors developing on a background of adiposity.

## METHODS

### Germline genetic data and BMI polygenic score

Blood-derived DNA (germline DNA) from the TCGA UCEC cohort had been genotyped by the TCGA Research Network using the Affymetrix Genome-Wide Human SNP Array 6.0. We obtained genetically inferred ancestry calls for this cohort from Carrot-Zhang, et al. based on a consensus of five independent ancestry classification methods (21). For the cases in the cohort classified as being of genetically inferred European ancestry we downloaded the genotypes called by the Birdseed algorithm from the controlled tier of the legacy Genomic Data Commons (GDC) portal accessed via an approved application to the database of Genotypes and Phenotypes (dbGAP). Genotypes with Birdseed confidence score >= 0.1 were set as missing (based on Affymetrix recommendation) and REF and ALT alleles were aligned to the forward strand. Ambiguous SNPs (SNPs with A/T or C/G alleles) were removed. Genotype calls with a SNP call rate >95%, minor allele frequency >0.5%, and Hardy-Weinberg equilibrium exact test P≥10^−6^; and samples with >95% call rate, inbreeding coefficient, F such that |F|<=0.2, and kinship coefficient <=0.0884 were retained. These filters were applied using VCFtools (v0.1.16) (22). Retained genotypes were phased and imputed to the 1000 Genomes Phase 3 (Version 5) reference panel (23) using Eagle2 (24) and Minimac4 (25), respectively.

Data for the BMI PGS were obtained from a GWAS meta-analysis that involved 434,794 adult women of European ancestry from the Genetic Investigation of Anthropometric Traits (GIANT) consortium and the UK Biobank (18). Specifically, we used effect size estimates (regression beta coefficients) and corresponding allelic information for index variants associated with BMI in women at genome-wide significance (P < 5×10^−9^) that were independent (*r*^2^ < 0.05) on linkage disequilibrium (LD) clumping performed using a reference panel of 20,275 unrelated individuals from the UK Biobank (18)). Each woman in the TCGA UCEC cohort was then assigned her genetically predicted BMI (BMI PGS) using the beta coefficients from the BMI GWAS meta-analysis with PGS calculation implement using the PRSice-2 software (26). SNPs with ambiguous alleles were removed and the final BMI PGS was calculated based on 242 SNPs captured in the GWAS and TCGA data. Specifically, for the PGS calculation, the betas for each BMI-associated index SNP were multiplied by the effect allele count for that SNP for each woman in the TCGA UCEC cohort. These numbers were then summed over all 242 SNPs to assign each woman in the cohort her BMI PGS. The PGS was standardized by subtracting the mean PGS for the cohort and dividing by the standard deviation.

### Tumor multi-omic data

An in-depth description, including details of biospecimen collection, laboratory protocols, quality control and bioinformatic processing pipelines, for the TCGA UCEC tumor multi-omic data sets is available in Levine, et al. (20). Here we briefly describe the specific data sets that were analyzed in our study. Tumor gene expression in the UCEC cohort was measured using the Illumina Genome Analyzer RNA-Seq platform. The “RNA-Seq by Expectation Maximization” (RSEM) raw count matrix including 20,531 genes was downloaded from the Firehose portal (gdac.broadinstitute.org). We removed the 10% of genes that had the lowest expression levels and subjected the matrix to quantile normalization and inverse normal transformation. Estimates of the relative fractions of 22 immune cell types within the leukocyte compartment of TCGA UCEC tumors were downloaded from the GDC (gdc.cancer.gov/about-data/publications/panimmune). These were derived from a previously published application of the “cell-type identification by estimating relative subsets of RNA transcripts” (CIBERSORT) algorithm to the UCEC RNA-Seq data (27, 28).

CIBERSORT uses a set of immune cell profiles as reference to calculate a baseline matrix of immune cell signatures that can then be applied to tumor samples to determine relative proportions of immune cells in the leukocyte fraction of the tumor. Tumor protein expression in the UCEC cohort was measured using the reverse phase protein array (RPPA) for 131 proteins and the “level 3” replicate-base normalized protein expression matrix was downloaded from the Xena repository (xena.ucsc.edu).

A matrix of 65 single-base substitution (SBS) tumor mutational signatures called from whole-exome sequencing of 9,493 tumors in the TCGA pan-cancer cohort was downloaded from the Xena repository and filtered to only retain the UCEC subset and mutational signatures with at least 10% prevalence in this subset (29). Tumor mutation load estimates (silent mutations per megabase (Mb) and non-silent mutations per Mb) linked to Thorsson, et al. (28) were also downloaded from the GDC for the pan-cancer TCGA cohort and subset to UCEC cases. “TCGA Unified Ensemble MC3” gene-level tumor mutation calls were downloaded from the Xena repository. These calls were obtained from a standardized variant calling pipeline involving an ensemble of seven mutation-calling algorithms with artefact filtering and variant annotation applied to the UCEC tumor exome sequencing data set and carried out as part of the Multi-Center Mutation Calling in Multiple Cancers (MC3) project (30). Genes in each TCGA tumor were subjected to a binary classification as either wild-type (zero) or carrying a non-silent mutation (one). We focused our analysis on 13 established cancer driver genes that were mutated in at least 10% of endometrial cancers nominated as per the IntegrativeOncoGenomics portal (intogen.org). Genome-wide DNA methylation in the UCEC cohort was measured using the Illumina Infinium HumanMethylation450 array. A matrix of the methylation beta values and corresponding array probe information mapped to cytosine-phosphate-guanine (CpG) sites in the human genome were downloaded from Xena. We removed 107,211/485,578 CpG sites with >80% missing data. Copy number variation (CNV) burden scores were downloaded from the GDC and comprised number of segments altered in the copy number profile for each UCEC sample and fraction of bases deviating from baseline ploidy (28, 31).

### Statistical analyses

We used generalized linear models (GLMs) adjusted for age and stage at diagnosis, 10 genetic ancestry principal components, and microsatellite stability status to evaluate the associations between the germline BMI PGS and TCGA UCEC tumor gene expression, immune signatures, protein expression, mutational signatures, and tumor mutation load estimates. The default GLM was linear regression but for immune and mutational signature and mutational load data we used quasi-Poisson GLMs. For mutational signatures with >70% zeros we fit zero-inflated negative binomial (ZINB) models. The GLMs for mutational signatures were also adjusted for signature accuracy. Microsatellite stability was coded as a binary variable (microsatellite stable (MSS) versus instability (MSI)). For the major non-null findings, we also re-evaluated the associations between BMI PGS and tumor molecular features by splitting the UCEC cohort into MSS versus MSI subgroups to assess relative strength of associations by microsatellite status. Age, stage, MSS/MSI covariates were obtained from the clinical phenotypes matrix file downloaded from Xena while principal components were obtained from Carrot-Zhang, et al (21).

In addition to examining gene expression associations at the level of single genes, we also performed pathway analysis. We ranked the 18,458 genes evaluated genome-wide in the BMI PGS-tumor gene expression analysis by descending order of their expression-association with the BMI PGS using the absolute value of the linear regression Z-score as the ranking metric. This ranked list was then subjected to gene set enrichment analysis (GSEA; (32)) to identify biological pathways enriched among genes whose tumor expression was strongly associated with the BMI PGS. The Molecular Signatures Database Hallmark gene set collection (33) comprising 50 pathways was used for GSEA (33). GSEA was implemented using fgsea (version 1.26.0) and msigdb (version 7.5.1) R packages. We also conducted an epigenome-wide association study (EWAS) using the CpGassoc R package (version 2.6; (34)) to test for associations between the germline BMI PGS and tumor DNA methylation in the UCEC cohort. The fixed-effects EWAS models were adjusted for the same covariates as the other GLMs described above. Finally, we examined associations between BMI PGS and four survival endpoints in the UCEC cohort: overall survival (OS), disease-specific survival (DSS), progression-free interval (PFI) and disease-free interval (DFI). The definitions and curation of these survival endpoints has been described previously in detail (35) and we downloaded the survival data from the Xena repository. Survival analyses were performed using Cox proportional-hazards models adjusted for age, stage, MSS/MSI status, and 10 genetic principal components and implemented using the survival (version 3.3.1) R package. For the top pathway-level gene expression and protein expression associations with BMI PGS, we used the cBioPortal to evaluate associations with OS, PFI, DSS and DFI using Kaplan-Meier plots and log-rank tests (36).

False discovery rate (FDR) control was used to account for multiple comparisons in each analysis described above and results at FDR<0.05 emphasized. To avoid over-reliance on a hard FDR threshold to guide our interpretation we also explored results achieving FDR<0.10 (37).

## RESULTS

### Correlation of BMI PGS and BMI measured at diagnosis

Of the 354 TCGA UCEC cases with matched tumor and germline data who were assigned a BMI PGS, height and weight at diagnosis were available and used to calculate the BMI at diagnosis for 337 patients. After removing 15 cases with extreme BMI values at diagnosis (BMI<15 kg/m^2^ or >50 kg/m^2^), the BMI PGS, which reflects germline genetically predicted BMI trajectory over the long term, was correlated (Pearson’s *r*=0.23; P=2.1×10^−5^; Supplementary Figure 1A) with the actual measured BMI at the timepoint of diagnosis (median BMI at diagnosis = 31.9 kg/m^2^, interquartile range (IQR) = 12.9 kg/m^2^). BMI at diagnosis is likely to be affected by the prior onset of the cancer itself and is a single-timepoint measure so we did not expect to see a perfect correlation with the BMI PGS nor complete independence either and the correlation of 0.23 reflects this trade-off. We also used linear regression to test for associations between the BMI PGS and age (median age = 64 years, IQR = 16 years) or stage (stages I/II/III/IV: 67%/8%/21%/4%) at diagnosis or tumor microsatellite status (MSS = 58%, rest MSI) but found no statistically significant associations with these variables (Supplementary Figure 1B-D). There was no visible patterning of the endometrial cancer histological and molecular subtypes by increasing BMI quintiles (Supplementary Figure 2A and 2B).

### Associations between BMI PGS and tumor gene expression

We tested for associations between the BMI germline PGS and the tumor RNA-Seq-based expression level of 18,458 genes across the genome in 349 TCGA UCEC cases with matched germline DNA-somatic RNA-Seq data. None of the single gene transcriptomic associations achieved FDR<0.05 after correcting for multiple comparisons (Supplementary Table 1). However, ranking the genes in descending order of their strength of tumor transcriptomic association with the BMI PGS and performing GSEA using the 50 Hallmark gene set biological pathways identified profound pathway-level enrichment, with tumor gene expression changes in 11 pathways associated with the BMI PGS at FDR<0.05 (Supplementary Table 2). The top pathway association with the BMI PGS was IL6-JAK-STAT3 signaling tumor expression (P=8.5×10^−9^; FDR=4.2×10^−7^; Figure 2; Supplementary Table 2). A total of 49 of the 86 genes in the IL6-JAK-STAT3 pathway were defined by GSEA as “leading edge” genes (i.e., responsible for driving the enrichment signal) and 44 of these 49 genes were upregulated with increasing BMI PGS (Figure 3A; Supplementary Table 3). These included both key drivers *IL6* (the gene most strongly upregulated with higher BMI PGS in this pathway; P=6.8×10^−4^; Figure 3B; Supplementary Table 3) and *STAT3* (the fourth most strongly upregulated gene; P=6.3×10^−3^; Figure 3C; Supplementary Table 3). The other pathways with FDR<0.05 highlighted a range of inflammatory and immune-mediated processes including allograft rejection and inflammatory and interferon gamma response pathways (Figure 2; Supplementary Table 2). Repeating the BMI PGS-tumor transcriptomic association and pathway analyses in subgroups defined by microsatellite stability status did not yield clear differences in pathway-level association by microsatellite status and the association with IL6-JAK-STAT3 signaling was observed in both subtypes (Supplementary Tables 4—7).

**Figure 1:**
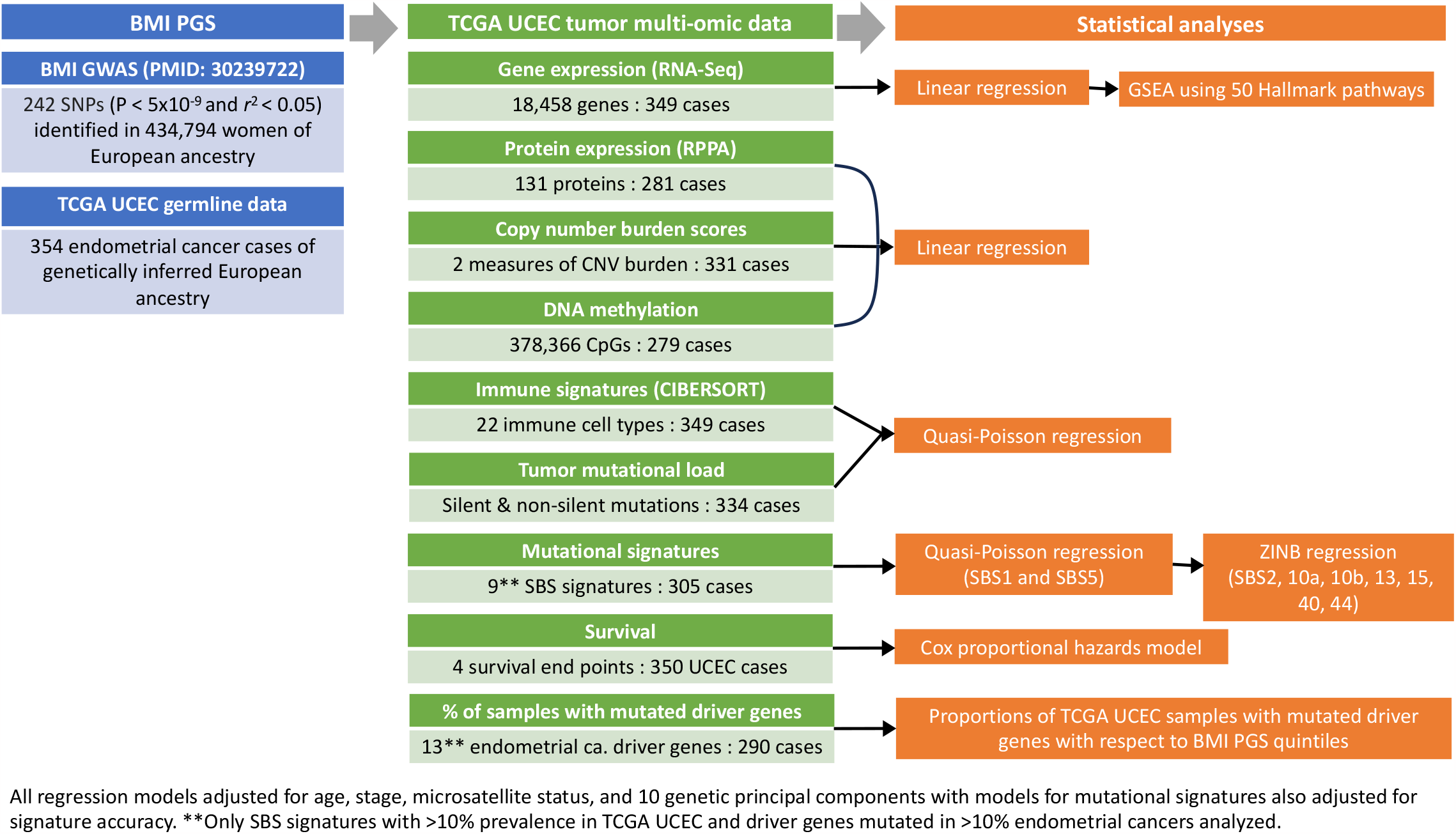
Schematic overview of study design, data sources, and analyses performed.

**Figure 2:**
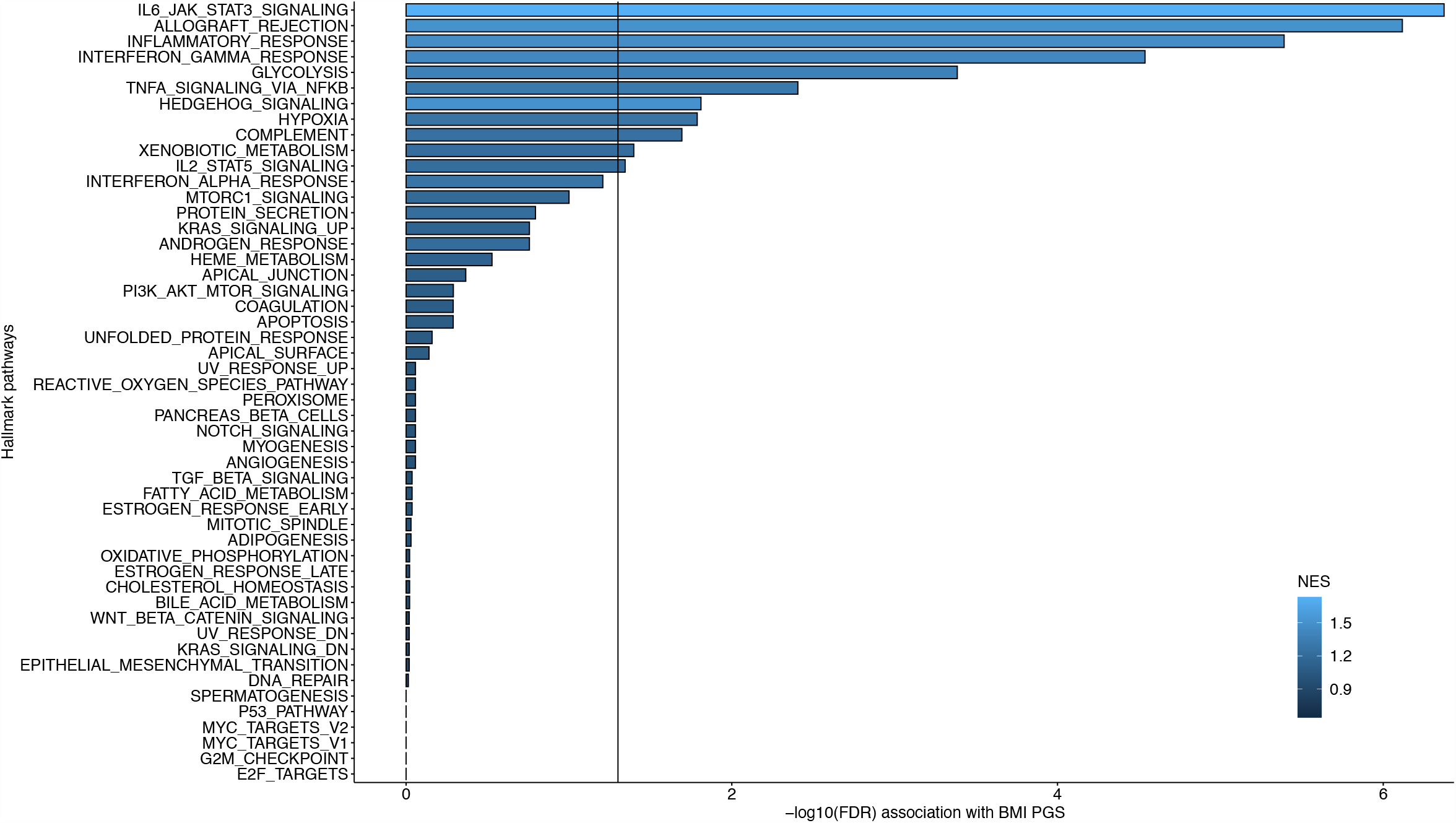
Gene set enrichment analysis (GSEA) results for 50 Hallmark pathways for the association between BMI PGS and tumor gene expression (RNA-Seq) in the TCGA UCEC cohort. The strength of enrichment associations corrected for multiple comparisons (-log_10_(FDR)) for each of the 50 pathway gene sets evaluated is shown. The vertical line indicates FDR = 0.05 and the histogram bars are colored based on GSEA normalized enrichment scores (NES) for each pathway.

**Figure 3:**
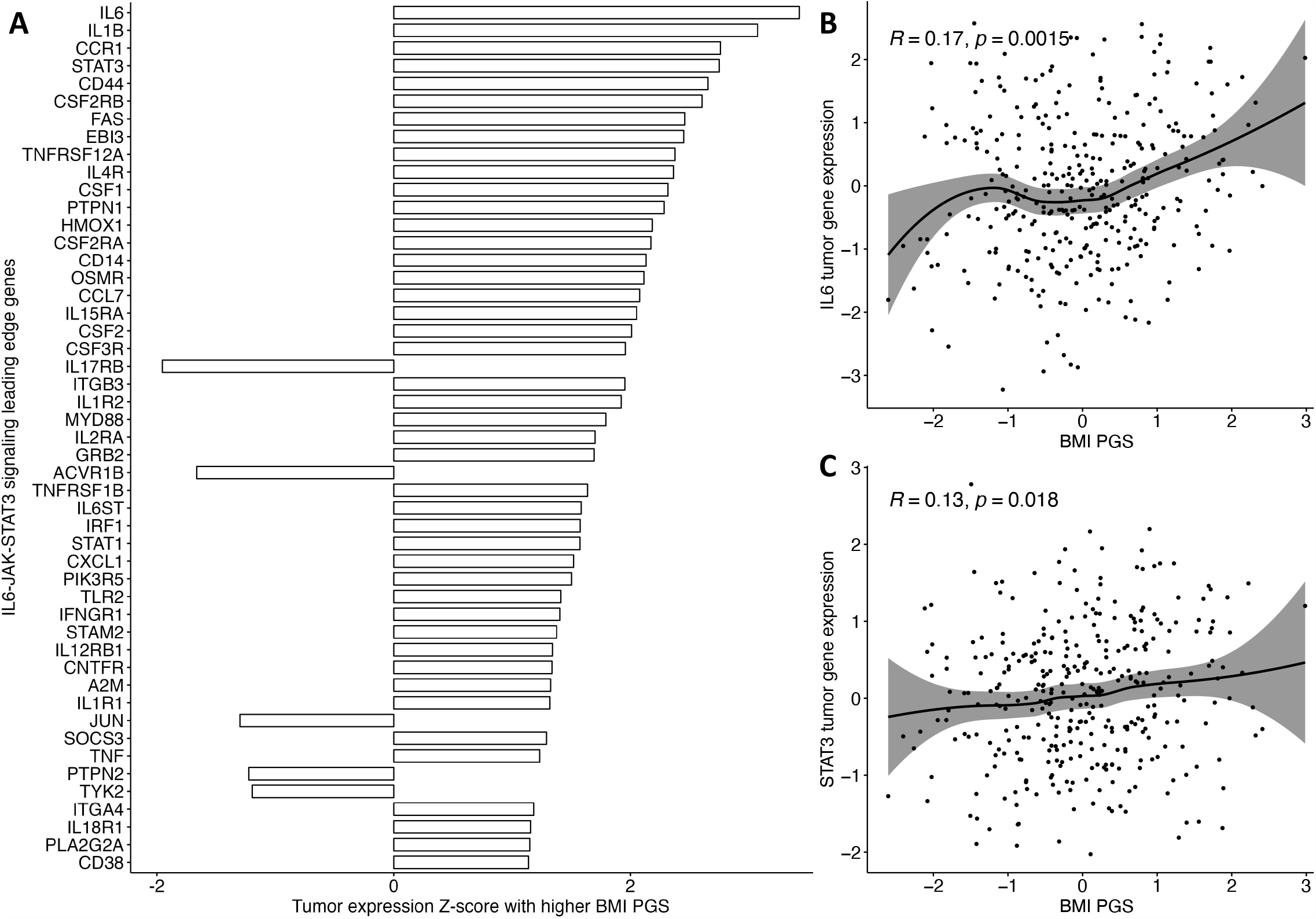
The direction of association (regulation) between BMI PGS and TCGA UCEC tumor expression of specific genes in the IL6-JAK-STAT3 signaling Hallmark pathway. (A) Z-scores with respect to higher BMI PGS for each of the 49 leading edge genes driving the IL6-JAK-STAT3 signaling pathway association in GSEA. Scatter plot and LOESS (locally weighted smoothing) line with 95% confidence interval shaded displaying the association between BMI PGS and inverse normal rank transformed tumor gene expression for (B) *IL6* and (C) *STAT3*. The Pearson correlation coefficient is shown while linear regression P-values adjusted for age, stage, microsatellite status, and 10 genetic principal components are provided in Supplementary Table 3.

### Associations between BMI PGS and tumor immune signatures

We examined associations between the BMI PGS and tumor CIBERSORT-based immune signatures that represented estimated intra-tumor infiltration levels of 22 immune cell types in the 349 TCGA UCEC cases with matched germline DNA-somatic RNA-Seq data. We found that higher BMI PGS was strongly associated with elevated levels of activated mast cell intra-tumor infiltration (Z-score=3.59; P=3.8×10^−4^; FDR=0.008; Figure 4A—C; Supplementary Table 8). The association was stronger in MSI tumors (Supplementary Table 9) than MSS tumors (Supplementary Table 10).

**Figure 4:**
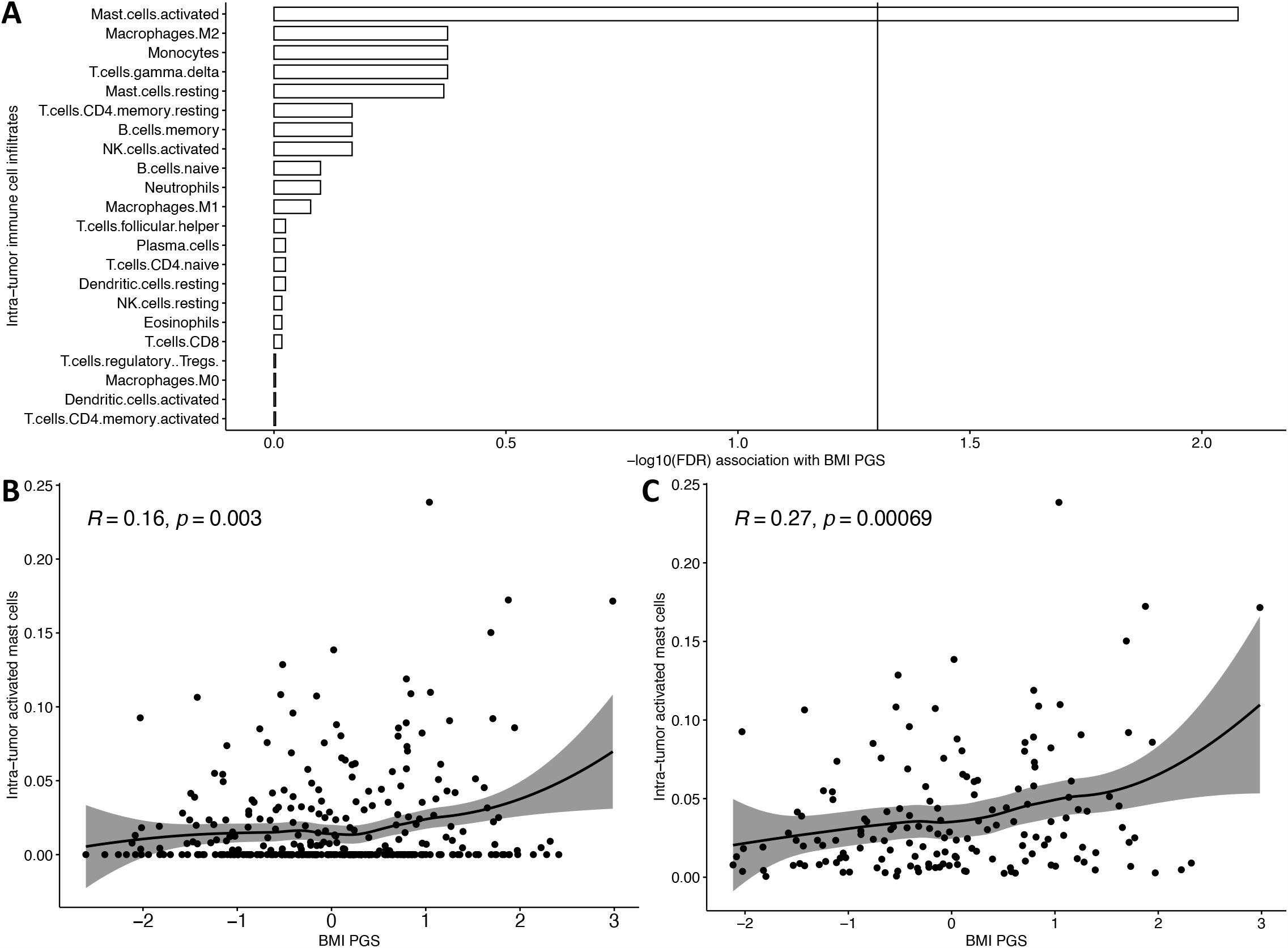
Associations between the BMI PGS and intra-tumor immune cell infiltration levels in the TCGA UCEC cohort. (A) Associations (-log_10_(FDR)) between the BMI PGS and the CIBERSORT-estimated intra-tumor infiltration levels for 22 immune cell types. The vertical line indicates FDR = 0.05. Scatter plot and LOESS (locally weighted smoothing) line with 95% confidence interval shaded displaying the association between BMI PGS and estimated intra-tumor activated mast cell infiltration levels in (B) the full TCGA UCEC cohort (N = 353) and (C) the TCGA UCEC cohort after removing tumors with a estimated activated mast cell infiltrate levels of zero (N = 161). The Pearson correlation coefficient is shown while linear regression P-values adjusted for age, stage, microsatellite status, and 10 genetic principal components are provided in Supplementary Table 8.

### Associations between BMI PGS and tumor protein expression

We evaluated associations between the BMI PGS and tumor RPPA-based protein expression levels in 281 TCGA UCEC cases with matched germline DNA-somatic RPPA data. The strongest association was observed for epidermal growth factor receptor (EGFR) expression, assayed by the EGFRPY1173 antibody, which was inversely associated with the BMI PGS (Z-score=-3.50; P=5.6×10^−4^; FDR=0.07; Figure 5 and 5B; Supplementary Table 11). This inverse association between tumor EGFR protein levels and BMI PGS was maintained in both MSS (Supplementary Table 12) and MSI (Supplementary Table 13) tumors.

**Figure 5:**
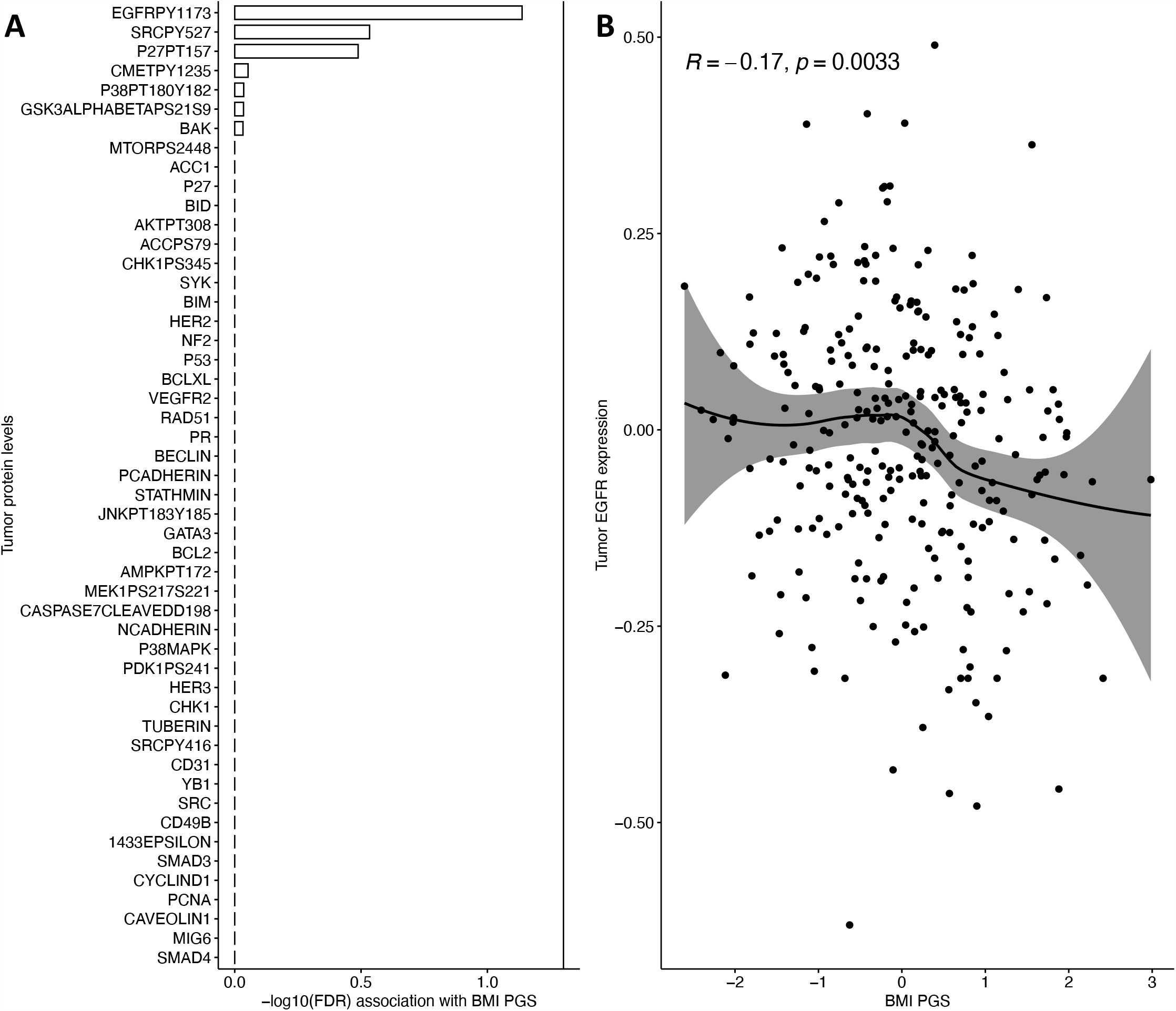
Associations between the BMI PGS and tumor protein expression in the TCGA UCEC cohort. (A) Associations between the BMI PGS and tumor protein expression for the top 50 proteins of the 131 assessed. The vertical line indicates FDR = 0.05. (B) Scatter plot and LOESS (locally weighted smoothing) line with 95% confidence interval shaded displaying the association between the BMI PGS and tumor EGFR protein levels. The Pearson correlation coefficient is shown while linear regression P-values adjusted for age, stage, microsatellite status, and 10 genetic principal components are provided in Supplementary Table 11.

### Associations between BMI PGS and tumor mutation data

We assessed the proportions of non-silent mutations by BMI PGS quintile for 13 somatic driver genes known to be frequently mutated in endometrial tumors using the 290 TCGA UCEC cases with matched germline DNA-somatic gene-level mutation data. We did not observe any clear patterns of increase or decrease in the proportion of mutations in each of these driver genes by progressively increasing quintiles of the BMI PGS (Figure 6A—M).

**Figure 6:**
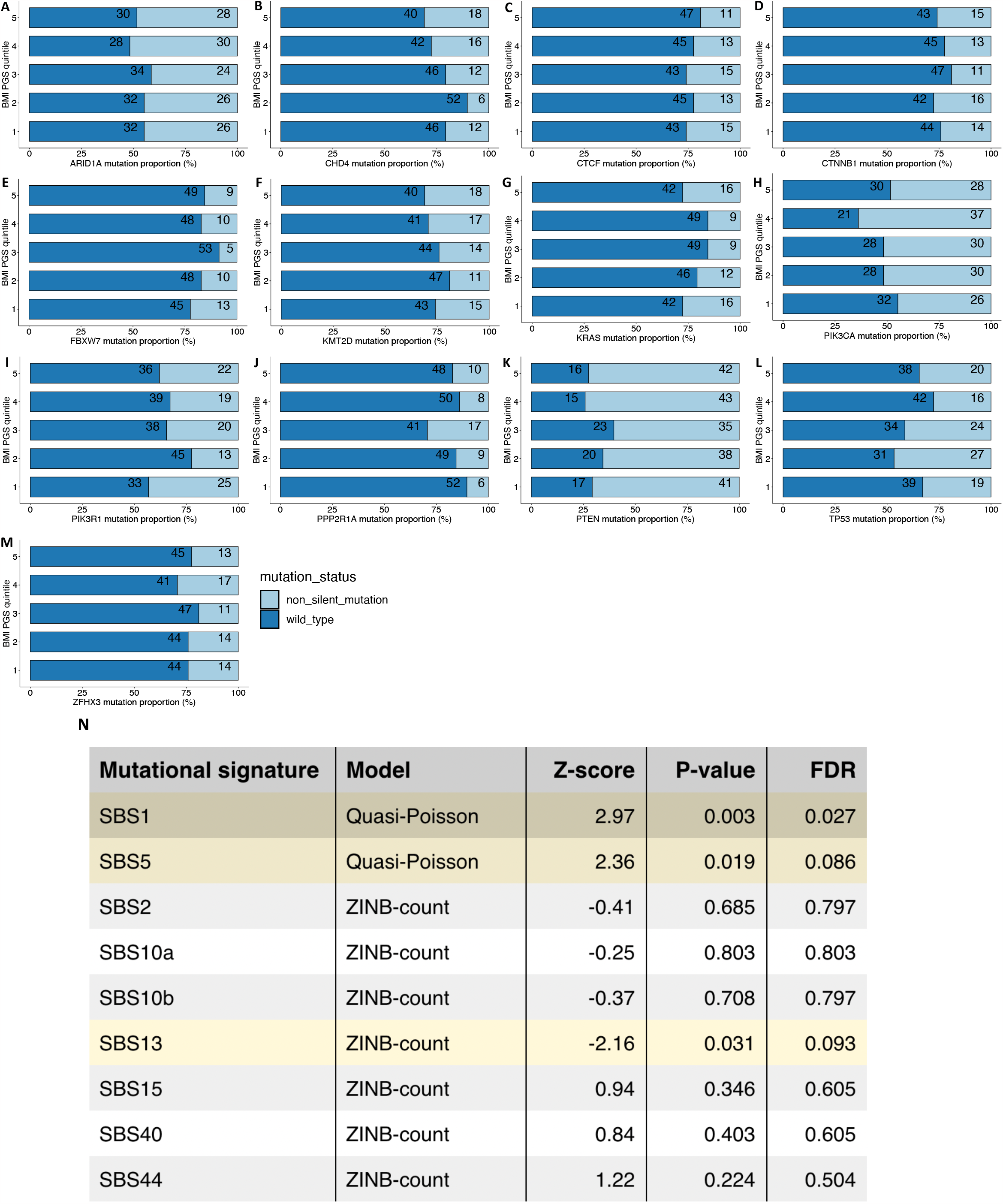
Tumor driver gene mutation status by BMI PGS quintile and BMI PGS-mutational signature associations in the TCGA UCEC cohort. Proportions of non-silent and wild-type mutations per BMI PGS quintile (increasing BMI PGS from quintiles 1 to 5) in 290 TCGA UCEC cases. We calculated these proportions for 13 somatic driver genes frequently mutated in endometrial cancers, including (A) *ARID1A*, (B) *CHD4*, (C) *CTCF*, (D) *CTNNB1*, (E) *FBXW7*, (F) *KMT2D*, (G) *KRAS*, (H) *PIK3CA*, (I) *PIK3R1*, (J) *PPP2R1A*, (K) *PTEN* (L), *TP53* and (M) *ZFHX3*. Mutational signature-BMI PGS associations (N) with associations achieving FDR < 0.10 highlighted.

Of the 65 single base substitution mutational signatures profiled in TCGA, SBS1 and SBS5 were observed with non-zero values in at least 94% of the UCEC cohort while SBS2, SBS10A, SBS10B, SBS13, SBS15, SBS40 and SBS44 had non-zero prevalence in 10%-28% of the cohort. Elevated BMI germline PGS was associated with higher levels of tumor SBS1 (Z-score=2.97; P=0.003; FDR=0.027) and SBS5 (Z-score=2.36; P=0.019; FDR=0.086) and lower SBS13 (Z-score=-2.16; P=0.031; FDR=0.093) across 305 UCEC cases with matched germline DNA-somatic mutational signature data (Figure 6N). In the subgroup analysis, these signature associations with the BMI PGS were more pronounced in the MSI than MSS subgroup (Supplementary Table 14). We found little evidence of association between the BMI PGS and levels of silent (Z-score=1.29; P=0.20) and non-silent (Z-score=1.13; P=0.26) mutations per megabase in 334 UCEC cases with matched germline DNA-tumor mutational load data.

### Associations between BMI PGS and tumor copy number burden scores, methylation, and survival

We found no evidence of associations between the BMI PGS and the number of segments (Z-score=0.14; P=0.89) and fraction of genome (Z-score=-1.15; P=0.25) subject to copy number alterations in the 331 UCEC cases with matched germline DNA-somatic copy number data. We also conducted an EWAS to identify associations between the BMI PGS and tumor DNA methylation levels across 378,366 genome-wide CpG sites in 279 UCEC cases with matched germline DNA-somatic methylation. None of the CpG sites reached epigenome-wide significance (P<1.3×10^−7^; Supplementary Table 15). We did not identify any statistically significant associations between the BMI PGS and the four survival endpoints (OS, DSS, DFI, PFI) in the 350 UCEC cases with matched PGS and survival data (Supplementary Table 16). However, aberrant expression in the UCEC cohort of the “leading edge” genes responsible for the IL6-JAK-STAT3 pathway-BMI PGS association was in turn associated with inferior survival (P_OS_=0.03 and P_PFI_=0.04, respectively; Figure 7A). Altered EGFR protein expression was also associated with poor survival (P_OS_=9.0×10^−3^ and P_PFI_=8.2×10^−3^; Figure 7B).

**Figure 7:**
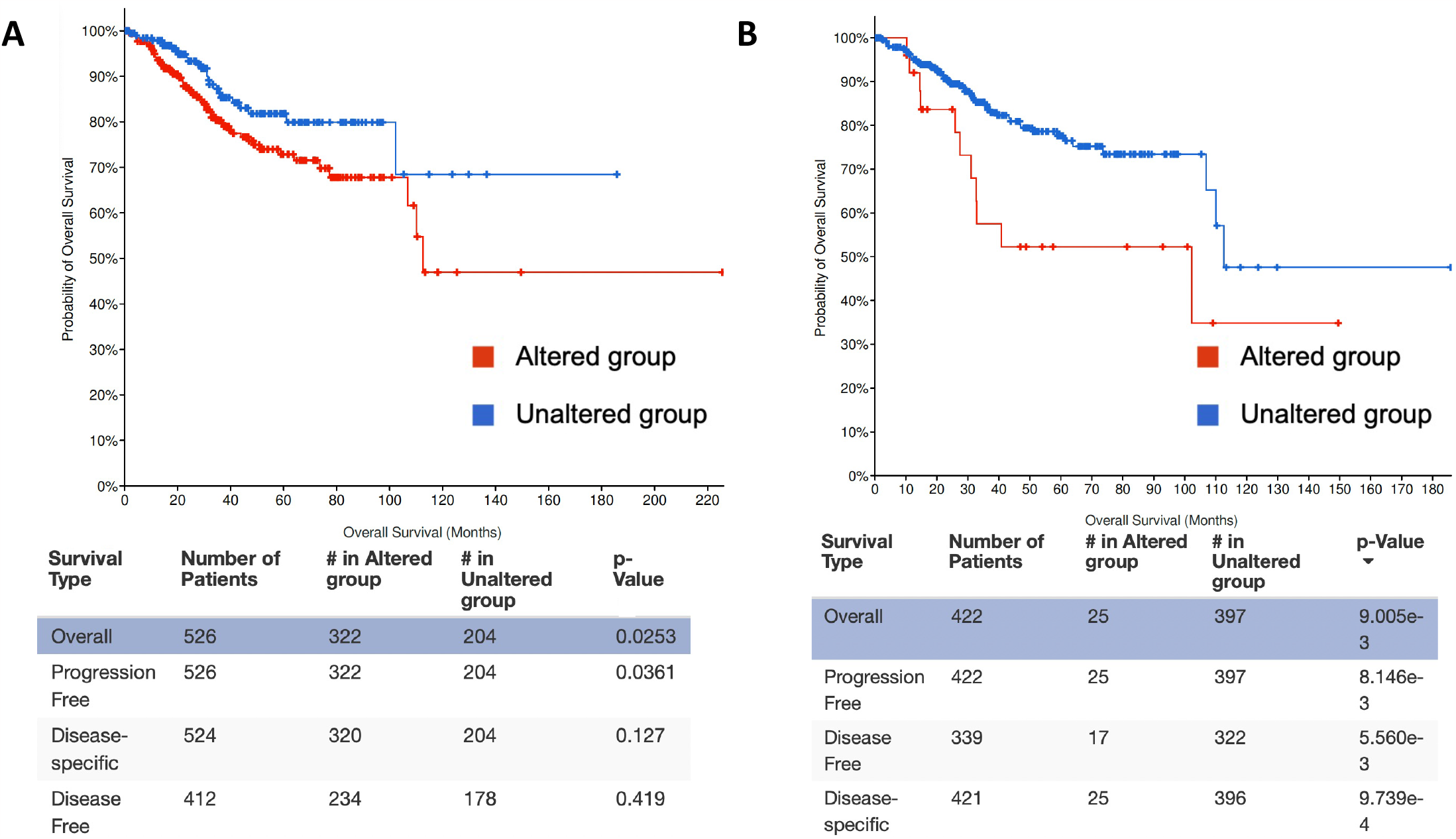
BMI PGS-associated tumor gene and protein expression features and their relationship with survival after a diagnosis of endometrial cancer in the TCGA UCEC cohort. (A) IL6-JAK-STAT3 pathway mRNA expression and overall survival, progression-free interval, disease-specific survival, and disease-free interval; with the Kaplan-Meier plot shown for overall survival and (B) EGFR protein expression and overall survival, progression-free interval, disease-specific survival, and disease-free interval; with the Kaplan-Meir plot shown for overall survival. P-values are from log-rank tests comparing TCGA UCEC cases with altered versus unaltered tumor gene or protein expression. Altered expression was defined as a Z-score more extreme than ±2.0. The Z-score for each case for a given gene/protein was computed by subtracting the mean expression calculated over the entire TCGA UCEC cohort from the expression in that case and dividing by the cohort-level standard deviation. IL6-JAK-STAT3 pathway membership for the survival analysis comprised 44 genes that were upregulated with higher BMI and drove the BMI PGS GSEA association for this pathway (i.e., the “leading edge” genes; Figure 3).

## DISCUSSION

Increased BMI is a causal risk factor for endometrial cancer. Despite this established association, the underlying molecular mechanisms and tissue-level pathways that link higher BMI to endometrial cancer development and the impact of long-term adiposity on endometrial tumor biology remain remarkably poorly understood. Against this background, here we adopt a unique approach examining associations between a germline PGS for BMI and tumor multi-omic data for endometrial cancer performing the largest and most comprehensive investigation of the effects of adiposity on endometrial cancer molecular profiles. The PGS represents the genetically predicted trajectory of BMI over the course of life that is more likely to capture the true long-term effects of adiposity on endometrial cancer initiation, development, and biology than BMI measured at any single time point such as on cancer diagnosis. Our study design was enabled by the availability of PGS from large-scale GWAS of adult BMI in women and blood-based germline genotype data matched to endometrial cancer tumor multi-omic data from TCGA. We identified associations between germline genetically predicted BMI and (i) tumor gene expression changes in multiple inflammatory and immune pathways, especially IL6-JAK-STAT3 signaling, (ii) intra-tumor mast cell infiltration, (iii) single base substitution mutational signatures 1, 5 and 13, and (iv) EGFR protein expression. We also show that BMI-linked aberrations in IL6-JAK-STAT3 pathway gene expression and EGFR protein expression having downstream impacts on survival after a diagnosis of endometrial cancer.

When we examined the association between germline genetically predicted BMI and tumor gene expression by placing them in the powerful biological context of well-defined molecular pathways, we identified associations between 11 pathways and genetically predicted BMI at FDR < 0.05, with the IL-6-JAK-STAT3 pathway yielding by far the strongest association. This suggests that BMI has profound effects on tumor transcription in endometrial cancers in a coordinated manner within these biological pathways. Notably, we observed widespread upregulation of genes involved in IL-6-JAK-STAT3 signaling, with *IL6* and *STAT3* ranking among the top four gene-level associations in this pathway. Obesity is increasingly being recognized as an activator of chronic JAK-STAT signaling mediated via cytokines such as IL6 (38). Aberrant hyperactivation of the IL6-JAK-STAT3 pathway is seen in several cancer types and associated with an inferior prognosis (39), and we found that that altered expression of genes in this pathway was also associated with worse survival in TCGA UCEC. Previous work in endometrial cancer cellular and mouse models has suggested that adipose-derived stem cells in the tumor microenvironment secrete IL6, which in turn activates its major downstream effector STAT3, and the two collectively promote endometrial cancer proliferation, invasion, and metastasis (40). Antibodies targeting the IL6 receptor and small-molecular inhibitors of JAK1 and STAT3 have been shown to inhibit endometrial cancer growth both *in vitro* in cell lines and *in vivo* in mice (41). The published literature also provides clues potentially implicating the obesity-driven pro-inflammatory interferon response in endometrial cancer pathogenesis, and in keeping with this we found that the other top ranked pathways significantly upregulated with higher genetic BMI in our analyses included the inflammatory response and interferon-gamma and -alpha pathways (42, 43).

We evaluated the association between germline genetically predicted BMI and the tumor infiltration levels of 22 immune cell types in the TCGA endometrial cancer cohort, identifying a significant association between higher BMI and elevated mast cell infiltrates. These infiltration levels were estimated using the CIBERSORT algorithm based on the tumor expression of genes that define these immune cell types, which has been shown to be a reasonable proxy for actual microscopically measured infiltration (44). Indeed, a previously published study involving microscopic analysis of 35 immunohistochemically stained surgically resected endometrial adenocarcinoma samples found that tumors with increased mast cell counts displayed greater myometrial invasion (45). Mast cell infiltration has been reported as an early feature in the development of various cancer types and interestingly mast cell release of IL-6 and other related angiogenic factors has been shown to have a vital role in neovascularization within the tumor microenvironment (46, 47). Tumor infiltrating mast cells are also known to be associated with resistance to anti-PD1 immunotherapy, which is increasingly being used to treat endometrial cancer (48). Activated mast cells express the proto-oncogene c-KIT, which encodes the target of tyrosine kinase inhibitors such as imatinib (47). Moreover, in a small series of 45 patients with endometrial adenocarcinoma, most tumors were found to express kinases that can be targeted by imatinib (49). Taken together with our findings, this suggests that mast cell infiltration in endometrial cancers developing on a genetic background of adiposity might represent a treatment target and calls for further research to explore this possible therapeutic vulnerability.

We identified significant associations between germline genetically predicted increased BMI and increased burdens of single base substitution mutational signatures SBS1 and SBS5, and decreased SBS13. Mutational signatures represent the genomic footprints of the impact of various exogeneous and endogenous cancer risk factors on the somatic or tumor tissues but the precise causes for most of the mutational signatures remain ill defined (50). SBS1 is characterized by C>T transitions at methylated CpG sites due to spontaneous deamination of 5-methylcytosine, DNA polymerase errors or enzymatic deamination. It is a “clock-like” mutational signature, but it is as yet unclear whether this clock represents time (aging) or the number of cell divisions. The exact molecular processes underlying SBS5 are unknown though it is also regarded as a “clock-like” signature correlated with age but not with cell division rate. SBS5 has been associated with chronic inflammatory states including inflammatory bowel disease and psoriasis (51, 52) and the association that we have uncovered between adiposity and SBS5 in endometrial cancer may reflect the consequence of an adiposity-driven low-grade chronic inflammatory state. Interestingly, SBS1 and SBS5 (along with SBS18, which we could not evaluate due to inadequate numbers) are the only SBS signatures found in normal endometrial epithelial cells (53). The prevalence of these signatures in normal endometrial glands was found to be correlated with age. Additionally, phylogenetic modeling indicated that the processes contributing to these signatures in normal endometrium were likely to remain active throughout life. This is consistent with recent multivariable Mendelian randomization analyses that have demonstrated that although adult adiposity has a greater impact on endometrial cancer risk, both childhood and adult adiposity remain associated with endometrial cancer risk (54, 55). Thus, placing our findings in the context of what is already known does suggest that obesity may well be a potential driver of SBS1 and SBS5 contributing to the development of endometrial cancer. SBS13 is associated with AID (activation-induced)/APOBEC (apolipoprotein B mRNA editing enzyme, catalytic polypeptide-like) cytidine deaminase DNA-editing activity and the inverse association with BMI PGS that we have uncovered suggests that this pathway may have a specific role in the development of endometrial cancer in those without the risk factor of adiposity.

We uncovered an inverse association at FDR<0.10 between germline genetically predicted BMI and endometrial cancer tumor EGFR protein expression and found that altered EGFR protein expression, in turn, was associated with a poor prognosis in TCGA UCEC. This suggests that endometrial cancers developing on a background with low levels of adiposity or that may lack adiposity as a risk factor exhibit higher levels of EGFR protein expression. Increased expression of the transmembrane epidermal growth factor receptor (EGFR) promotes cell growth, proliferation, and survival and is frequently observed in many cancer types (56). This association mirrors the inverse pattern observed in lung cancers, where non-smokers have a higher likelihood of carrying EGFR mutations that result in EGFR overexpression, compared to smokers with lung adenocarcinoma (57-59). Phase II clinical trials evaluating the EGFR inhibitor erlotinib (60) and more recently cetuximab (61) in recurrent or metastatic endometrial cancer have achieved promising response rates but failed to find specific molecular markers of this response such as EGFR mutation or copy number status. While requiring substantial additional work, we speculate that our results may inform the therapeutic targeting of EGFR in endometrial cancer by narrowing the focus of clinical investigation specifically to women who develop this cancer without a history of obesity.

Our study has some limitations. First, our analyses were restricted to individuals of European ancestry, and it is unclear whether our findings are generalizable to other populations. The restriction was enforced by the fact that most women in the TCGA UCEC and the BMI GWAS cohorts were of predominantly European genetic ancestry. Second, while we report several statistically robust associations between germline genetically predicted BMI and tumor multi-omic traits in the TCGA UCEC cohort, we note that there is an acute paucity of endometrial cancer data sets with matched germline genotype and multi-omic tumor phenotyping of comparable depth, breadth, and size to TCGA. The generation of additional and ancestrally diverse endometrial cancer data sets with matched germline and somatic profiles is warranted to further validate our findings. Third, unopposed estrogen signaling is a major risk factor for endometrial cancer and we did not attempt to jointly model the interaction between BMI and lifelong genetically predicted estrogen levels in our study design (62). Given the novel study design implemented here, investigating associations between a germline genetically predicted risk factor (BMI) and tumor molecular features, including a second risk factor in a multivariable or factorial setting would have made interpretation more complex. In sum, we cannot rule out the possibility that some of the BMI-tumor trait associations presented here are mediated via estrogen or other hormones such as insulin but that should not detract from the fact that these are ultimately driven by adiposity. Fourth, associations from case-only analyses such as ours are susceptible to collider bias. For example, if another endometrial cancer risk factor but not BMI is positively associated with an endometrial cancer tumor molecular trait, an endometrial cancer case developing on a background of high BMI may have lower levels of the other risk factor and therefore lower levels of the tumor molecular trait inducing a spurious and inverse association with high BMI. Ideally, our case-only findings should be replicated in a pre-cancerous endometrial tissue setting to ensure that these associations are operating over the longitudinal process of tumor development. However, such longitudinal normal or pre-cancer tissue collections with germline and multi-omic profiling are not currently available to our knowledge and addressing their lack has been highlighted as a priority for the field (63).

In conclusion, we identified associations between BMI predicted by germline genetic variants and endometrial cancer tumor gene expression changes in several immune and inflammatory pathways, most notably the IL6-JAK-STAT3 pathway; tumor mast cell infiltration estimates; mutational signatures (SBS1, SBS5, and SBS13); and tumor EGFR protein expression. Though requiring replication in independent endometrial cancer and pre-cancer cohorts with integrated germline-somatic data, these associations offer fresh and potential clinically actionable insights into the impact of adiposity on endometrial cancer biology.

## Data Availability

All data referred to in the manuscript are available at https://www.cancer.gov/tcga, https://gdac.broadinstitute.org/, https://gdc.cancer.gov/about-data/publications/panimmune, https://xena.ucsc.edu/, https://portal.gdc.cancer.gov/, and https://github.com/lindgrengroup/fatdistnGWAS/blob/master/SuppTable1/bmi.giant-ukbb.meta.1.merged.indexSnps.females.parsed.txt

https://gdac.broadinstitute.org/

https://gdc.cancer.gov/about-data/publications/panimmune

https://xena.ucsc.edu/

https://portal.gdc.cancer.gov/

https://github.com/lindgrengroup/fatdistnGWAS/blob/master/SuppTable1/bmi.giant-ukbb.meta.1.merged.indexSnps.females.parsed.txt

## ACKNOWLEDGMENTS

GR and AF are supported by Cancer Research UK [grant number C18281/A30905]. VG and SPK are supported by a UK Research and Innovation Future Leaders Fellowship to SPK [grant number MR/T043202/1]. KL, JMS, and SPK also receive support from the US National Institutes of Health (grant numbers R01CA211574 and R01CA259058). TR is supported by a UK National Institute for Health and Care Research (NIHR) Development and Skills Enhancement Award (grant number NIHR302363). EJC is supported by a NIHR Advanced Fellowship (grant number NIHR300650) and the NIHR Manchester Biomedical Research Centre (grant number NIHR203308). TRG is supported by the Medical Research Council Integrative Epidemiology Unit (grant number MC_UU_00032/03). EEV is supported by Diabetes UK [grant number 17/0005587] and the World Cancer Research Fund (WCRF, UK), as part of the WCRF International Grant Programme [grant number IIG_2019_2009]. TR, TRG, CLR and EEV are supported by Cancer Research UK [grant number C18281/A29019]. The results published here are in part based upon data generated by the TCGA Research Network: https://www.cancer.gov/tcga.

