## Supplementary Figures 1 and 2 for "The tumor multi-omic landscape of endometrial cancers developed on a germline genetic background of adiposity"

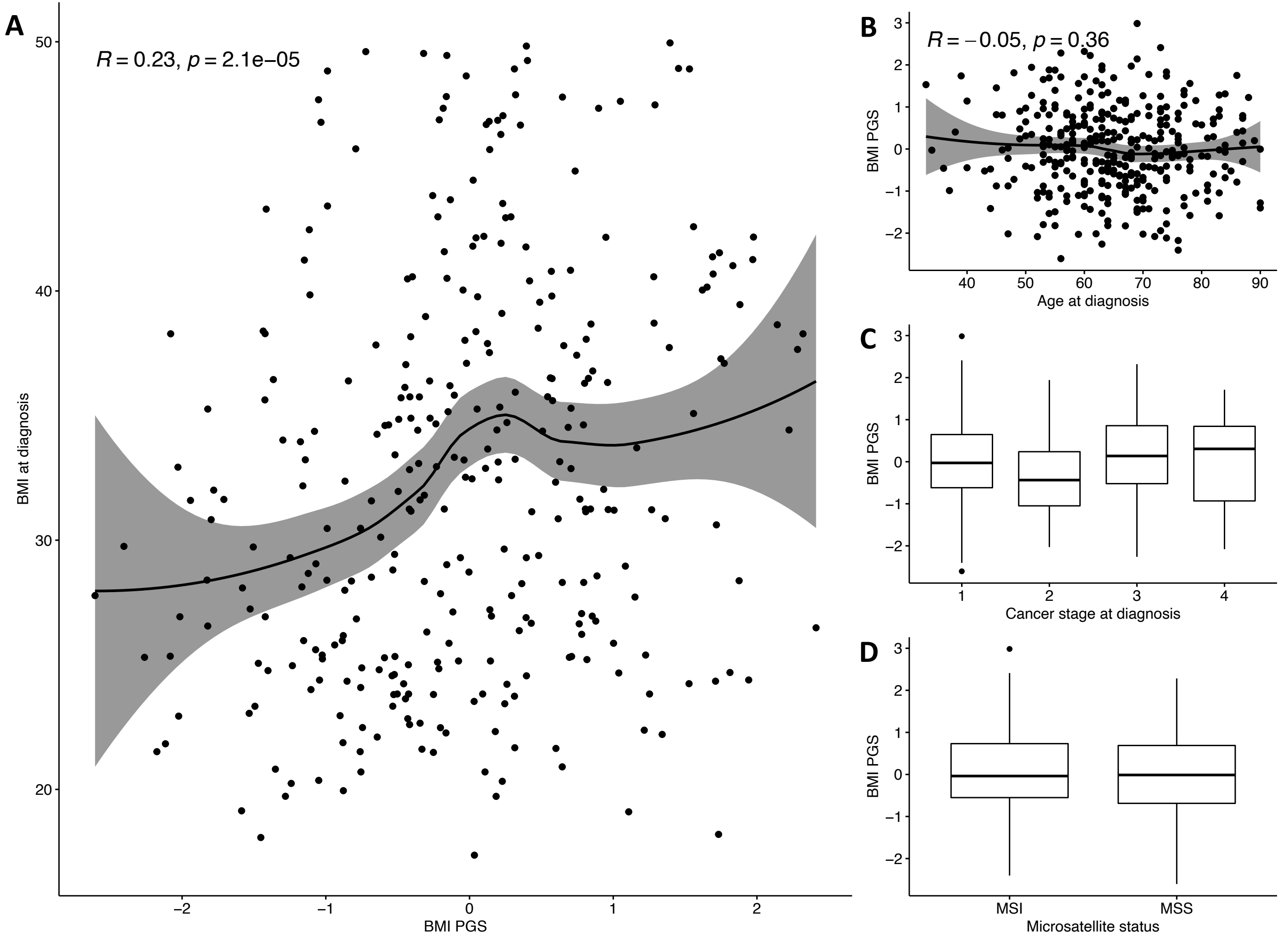


**Supplementary Figure 1:** Associations between the BMI germline PGS and (A) BMI measured at diagnosis (Pearson correlation coefficient; N = 322), (B) age at diagnosis (Pearson correlation coefficient; N = 354), (C) stage at diagnosis (N = 354; simple linear regression beta = 0.05 / P = 0.39), and (D) tumor microsatellite status (N = 351; simple linear regression beta = 0.04 / P = 0.72). The scatter plots in (A) and (B) include the LOESS (locally weighted smoothing) line with 95% confidence interval shaded.

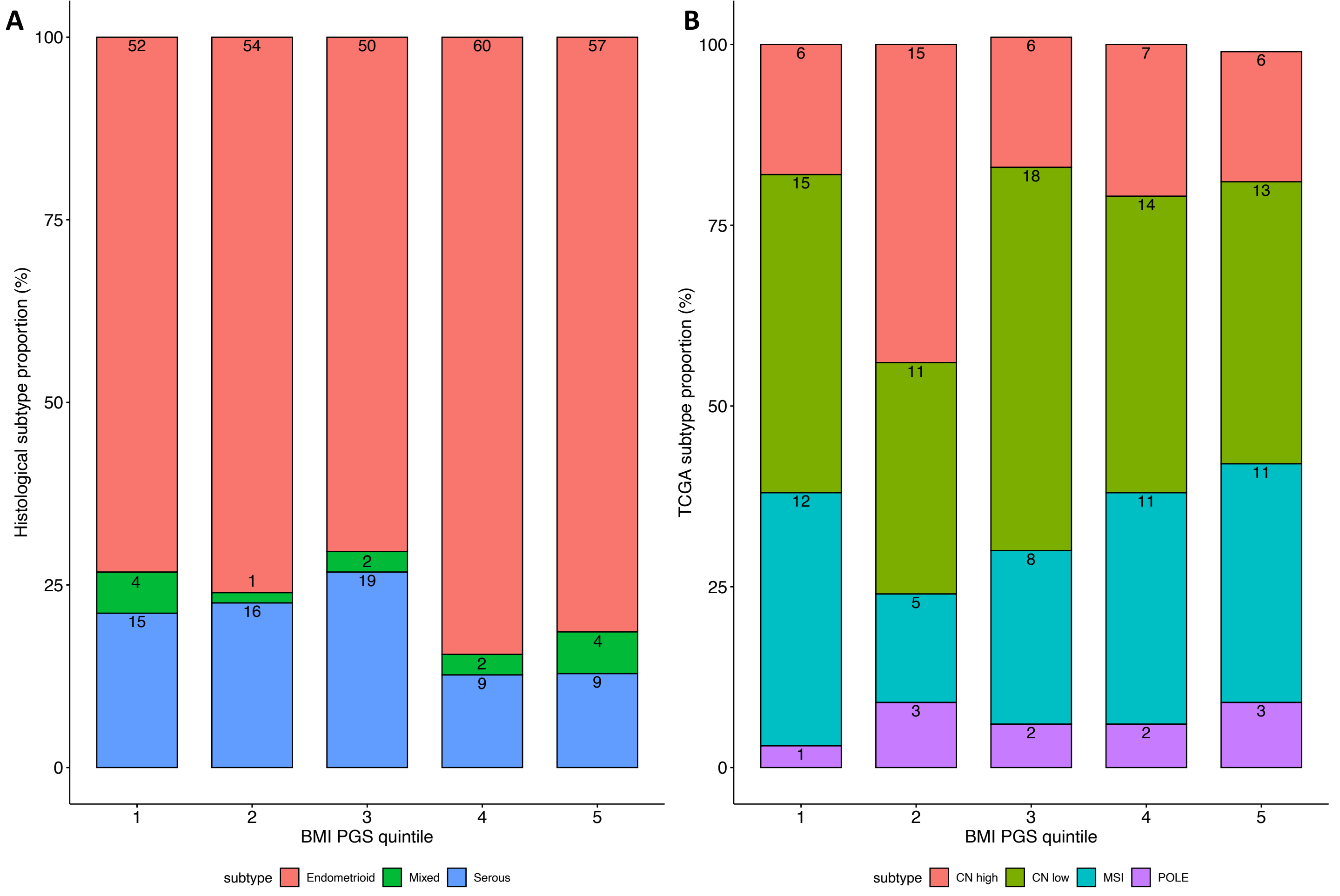


**Supplementary Figure 2:** Distribution of endometrial cancer histological (A) and TCGA molecular (B) subtypes by BMI germline PGS quintiles in the TCGA UCEC cohort.
